# White matter lesion volumes on 3-T MRI in people with MS who had followed a diet- and lifestyle program for more than 10 years

**DOI:** 10.1101/2024.04.04.24305252

**Authors:** Mariaan Jaftha, Frances Robertson, Susan J van Rensburg, Martin Kidd, Ronald van Toorn, Merlisa C. Kemp, Clint Johannes, Kelebogile E. Moremi, Lindiwe Whati, Maritha J Kotze, Penelope Engel-Hills

## Abstract

**Background:** Cerebral white matter lesions (WMLs) in people with multiple sclerosis (pwMS) are associated with the death of myelin-producing oligodendrocytes. MS treatment strategies aim to limit WML accumulation and disability progression. It is commonly accepted that nutrition is one of the possible environmental factors involved in the pathogenesis of MS, but its role as a complementary MS treatment is unclear and, to a large extent, ignored.

**Objective:** A pilot case control study over a 10 year period to ascertain whether a dietary- and lifestyle modification Program in pwMS reduces or prevents WML formation.

**Methods:** MRI was performed at baseline and after an interval period of at least 10 years or longer in 22 pwMS. WML volumes were determined using Sequence Adaptive Multimodal SEGmentation (SAMSEG) software, part of FreeSurfer 7.2. Other variables include age at MRI, disease duration, disability status and medication.

**Results:** PwMS (n=13) who had followed the Program for more than 10 years, had significantly smaller lesion volumes (mm^3^) compared to pwMS who did not adhere to the Program (n=9) (4950 ± 5303 vs 17934 ± 11139; p=0.002). WML volumes were significantly associated (p=0.02) with disability (EDSS) but not with age (p=0.350), disease duration (p=0.709), or Interferon-β treatment (p=0.70).

**Conclusion:** Dietary- and lifestyle changes may lower the risk of developing cerebral WMLs in pwMS and potentially slow down disease progression. Larger studies are required to confirm the effectiveness of such interventions in pwMS.

## Introduction

Multiple sclerosis (MS), the most common inflammatory disorder of the central nervous system (CNS) affecting young adults, is associated with damage to the myelin sheaths surrounding the nerve axons in the CNS. This causes disruption of signal transmission within the CNS and from the CNS to the peripheral organs, resulting in disability (loss of function). Radiologically, this manifests as cerebral white matter lesions (WMLs).^1^ Neuropathological studies confirm that early WMLs are associated with loss of myelin *secondary* to the death of myelin-producing oligodendrocytes.^1^ The precise mechanism of cell death and phagocytosis of myelin are still unclear as both activated microglia (local macrophages) and monocyte-derived macrophages from the circulation remove dead oligodendrocytes and dysfunctional myelin,^2^ while T-cells are rare and plasma cells (B-cells) are absent in these lesions. ^1,2^ Concomitantly, oligodendrocyte precursor cells (OPCs) migrate to the site of the lesion and remyelinate the axons.^1^ (Figure 1). OPCs are stem cells originating in the ventricular zones that remain abundant in the CNS, generating myelinating oligodendrocytes throughout adult life. ^3^ Therefore, if the remyelination is sufficient, the loss of function experienced by people with MS (pwMS) may be transient.

**Figure 1.**
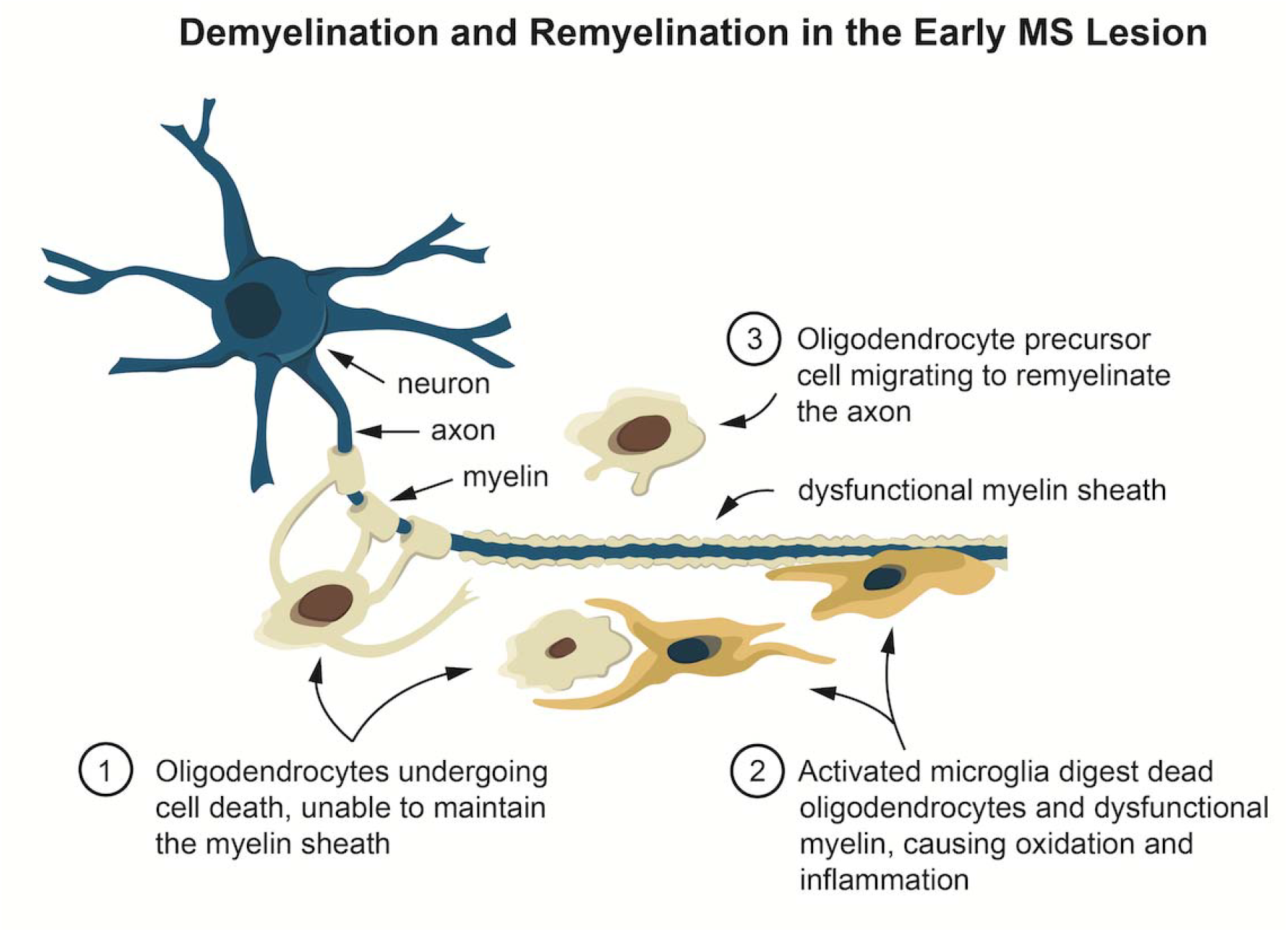
Demyelination and remyelination of the early MS lesion as revealed by neuropathological studies [1,2]. Oligodendrocytes experience cell death and are unable to maintain the myelin sheath, then activated microglia or monocyte-derived macrophages digest (scavenge) the dead oligodendrocytes and dysfunctional myelin, causing white matter damage and disability. Thereafter oligodendrocyte precursor cells (OPCs) become the new oligodendrocytes and remyelinate the axons, restoring functionality.

The reason(s) for oligodendrocyte demise and consequent MS disability has been the ongoing subject of a cross-sectional research study approved by the University of Stellenbosch Ethics Committee in 1996.^4,5^ This has involved pathology-supported genetic testing (PSGT) to search for genetic and environmental risk factors for demyelination.^6,7^ Following a study that recruited 118 pwMS to investigate the role of iron in MS,^8^ subsequent sub-studies found that MS risk factors fall into two categories: (1) *deficiencies*: (iron, vitamin D, vitamin B12, antioxidants, unsaturated oils, decreased CNS blood flow) and (2) *aggravators*: (oxidation/inflammation, smoking, infections, dietary saturated fat, increased cholesterol/homocysteine/obesity; allergies/food sensitivity and psychological stress). The rationale was outlined previously ^5,6,7^ and in the Discussion below.

Since pwMS are not all affected similarly, they are offered the opportunity to enrol at our institution in a PSGT Program that allows assessment of their personal risk factors and optimization of cerebral nutrients. The program entails performance of biochemical tests that allow for the identification of possible deficiencies of iron, vitamin D and vitamin B12, or excess cholesterol/homocysteine ratios and increased inflammation (CRP). This is complemented by genetic testing aimed at identification of clinically relevant variants in metabolic pathways and integration thereof with family history, biochemistry and lifestyle data to generate personalized reports.^5,9^ These reports enable clinicians to address imbalances revealed in the blood test investigations so that all the biochemical risk factors are mitigated through nutritional supplementation and modification of diet. In addition, the provided reports address lifestyle risks such as smoking and lack of exercise which potentially impair cerebral circulation, which is vital for adequate delivery of oxygen and nutrients to the oligodendrocytes.^10,11^ Details about the PSGT program and its beneficial effects have been comprehensively described in several publications.^5,6,7^

It is commonly accepted that nutrition is one of the possible environmental factors involved in the pathogenesis of MS, but its role as a complementary MS treatment is unclear and, to a large extent, ignored. Nutrition in MS has increasingly generated interest since the studies of Swank in the 1950’s which suggested that dietary saturated fat intake was associated with prevalence and disability in MS.^12^ Wider acceptance by the medical profession has however been limited by the fact that Swank’s studies were observational and not randomized controlled trials (RCTs). That problem has now been addressed: a Systematic Review and Network Meta-analysis of Randomized Trials by Snetselaar et al. (2023) has included 12 RCTs comparing 8 dietary interventions (low-fat, Mediterranean, ketogenic, anti-inflammatory, Paleolithic, fasting, calorie restriction, and control [usual diet]).^13^ The Wahls modified Paleolithic and Swank low fat diets both significantly improved quality of life (QOL) and fatigue,^14^ while the Mediterranean diet was significantly associated with significantly lower disability in pwMS.^15^ An RCT that excluded both saturated fat and unsaturated oils such as olive oil, found an improvement in fatigue and QOL but not in disability or MRI outcome measures.^16^ In contrast, WMLs were shown to be significantly decreased in some brain regions in pwMS who followed an anti-inflammatory diet.^17^ A limitation of the published diets for MS is that their benefits for preventing disability progression may be overruled by other risk factors, such as smoking (active or passive).

The present proof-of concept pilot study tested the hypothesis that relapse-associated worsening (RAW) and WML accumulation could be delayed, and serious MS disease prevented, if all the risk factors for oligodendrocyte demise mentioned above are addressed. The hypothesis was tested using 3-Tesla MRI to measure WML volumes and disability assessments together with diet- and lifestyle information.

## Methods

### Study participants

This study formed part of a case-control sub-study that recruited 51 pwMS (48 females and 3 males) and 25 Controls without neurological symptoms, for vascular ultrasound of the carotid arteries^10^ and assessment of disability using the Expanded Disability Status Scale (EDSS), as well as genetic studies.^5^ Twenty-five of the female pwMS as well as 25 age-matched female Controls who had demographic, biochemical and lifestyle data, were randomly recruited for 3-Tesla (3T) MRI at the Cape Universities Body Imaging Centre (CUBIC), University of Cape Town. Of these, 22 pwMS and 21 Controls completed the MRI.

PwMS were diagnosed before study entry according to the McDonald criteria (2001).^18^ Controls without neurological disorders were age- and sex matched. Exclusion criteria were a BMI > 30, any factors contrary to having an MRI scan, smoking, diabetes, or previous cardiac/neurological conditions. Demographic data recorded were age at MRI, disease duration (years with MS), and medication. Disease modifying treatment (DMT) was Interferon-β in 10 of the pwMS. The pwMS voluntarily decided whether to implement the recommendations to follow the PSGT Program or not. Of the 22 pwMS, 13 had followed the Program for more than 10 years, while 9 had not followed the Program.

### Disability assessments

Disability was assessed by a clinician using the EDSS to quantify disability in the pwMS.^19^ It is widely used to monitor changes in the level of disability in pwMS. The EDSS scale ranges from 0 to 10 in 0.5 unit increments that represent higher levels of disability. Scoring in the study was based on an examination by a neurologist.

### MRI procedures and software analysis

Of the 25 PwMS and 25 Controls randomly recruited from the vascular ultrasound study, 7 (3 pwMS and 4 Controls) did not have an MRI due to concerns of claustrophobia or illness. In total, 22 females with MS and 21 female Controls signed consent to be included in the 3-T MRI assessment.

Participant preparation occurred outside of the scan room and this process included an explanation of the procedure to ensure that the participant was comfortable as well as a CUBIC MRI compatibility checklist, signed by both the participant and the radiographer before removing their clothing/jewellery, etc., and putting on a cotton gown. Scanning proceeded in the head-first, supine position. A pillow was placed under the head and a wedge under the knees of each participant to lessen their discomfort during the scan. To ensure privacy, participants were covered with a sheet or a light blanket. A panic ball was handed to all participants to be used as an alarm in case they required any assistance during the procedure. MRI-compatible headphones were placed over their ears to reduce the acoustic noise of the scanner and to ensure participants could hear instructions during the scan.

Images were acquired on a Siemens Magnetom Skyra scanner using a standard 32-channel transmit receive (RF) head coil. The protocol included the following sequences: Fluid Attenuation Recovery (FLAIR) in axial and sagittal planes, Proton Density (PD), T2 (in coronal plane), Magnetization Prepared Rapid Gradient Echo (MPRAGE),Susceptibility Weighted Imaging (SWI), Diffusion Tensor Imaging (DTI), and Readout SEgmentation Of Long Variable Echo-trains (Resolve) images. The axial FLAIR and T1 sequences were specifically employed for the data analyses to provide contrast and expose pathology. Imaging parameters for these sequences are shown in Table 1.

**Table 1:**
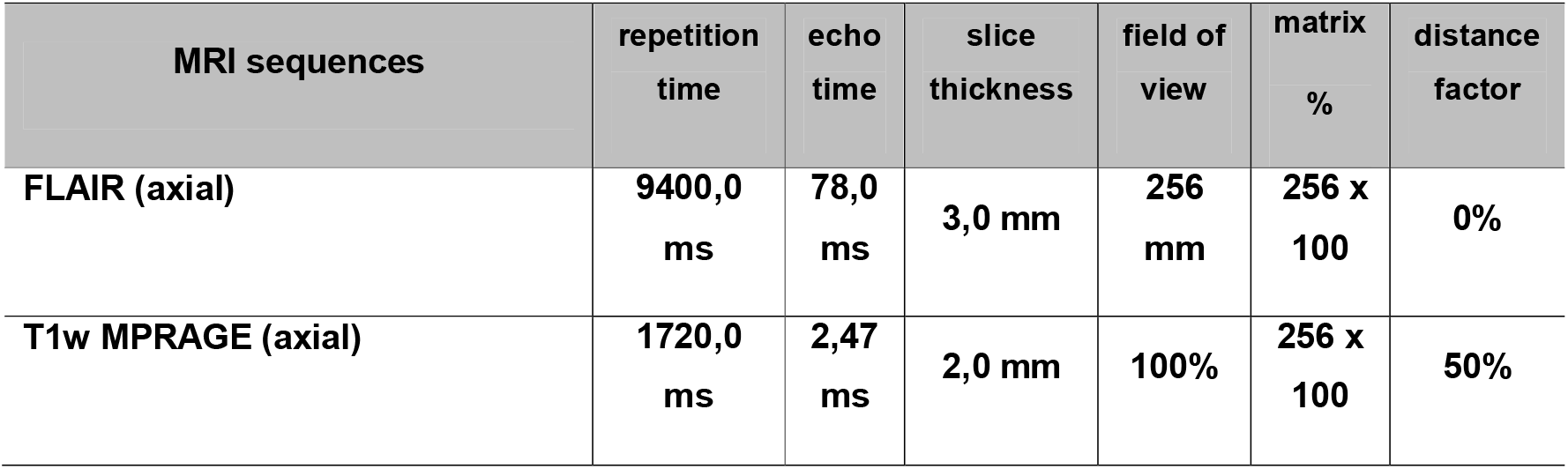
MRI Sequences and Parameters.

Lesion volumes were determined using the FLAIR and T1 images with Sequence Adaptive Multimodal SEGmentation (SAMSEG) software,^20^ part of FreeSurfer 7.2.

Data were generated to characterise lesion load, disability, demographic and biochemical markers.

### Biochemical analysis

Blood was drawn for biochemistry testing between 09h00 and 10h30 to standardize for diurnal variation. Biochemical analysis of the following biomarkers was performed by an accredited pathology laboratory: iron parameters, ferritin, C-reactive protein (CRP), total cholesterol, homocysteine, serum folate, vitamin B12 and 25-OH vitamin D as reported previously.^9^

### Statistical analysis

Statistical analyses were performed using Statistica™ version 13.4.0.14. Differences in lesion volumes and EDSS parameters between study groups were tested using one way analysis of variance. Normality was assessed by inspecting normal probability plots, and were in all cases found to be acceptable. Levene’s test was done to test for homogeneity of variance, and was in all cases accepted. In cases where it did not hold, the Welch test was also done. However the outcome of the Welch test was the same as for the F-test of the ANOVA, so only the ANOVA result was reported. Pearson correlations were reported to test for relationships between lesion volumes, lifestyle parameters and EDSS.

A 5% significance level was used as guideline for statistical significance.

### Ethics

This study was a collaborative investigation between Stellenbosch University (SU), the University of Cape Town (UCT) and the Cape Peninsula University of Technology (CPUT) in the Western Cape, South Africa. Ethics approval was granted by the Faculty of Medicine and Health Sciences Research Ethics Committee of the SU (references NO7/09/203 and N09/08/224) and the Faculty of Health and Wellness Sciences Research Ethics Committee of CPUT (reference no: CPUT/HW-REC 2019/H27). As a postgraduate project the study was conducted according to the code of ethics of the World Medical Association.^21^ All study participants gave signed informed consent for the MRI and for their data to be used for a research publication.

## Results

The clinical data of 22 pwMS and 21 Controls are summarized in Table 2.

**Table 2.** Clinical data of 22 pwMS and 21 Controls who had 3-T MRI assessments.

| Parameter | PwMS on PSGT Program for >10 years |  | All pwMS (n=22) | Controls (n=21) |
| --- | --- | --- | --- | --- |
|  | YES | NO |  |  |
| Females n | 13 | 9 | 22 | 21 |
| Age at MRI: years, mean (range) | 52 (36-65) | 48.8 (23-71) | 50.7 (23-71) | 48.3 (25-72) |
| Disease duration: years, mean (range) | 17.5 (11-29) | 12.2 (1-27) | 15.4 (1-29) | NA |
| Disease modifying therapy | 3 | 7 | 10 | NA |
| Untreated n (%) | 10 | 2 | 12 | NA |
| BMI kg/m<sup>2</sup>, mean (range) | 25.2 (21.5-35) | 25.5 (19-32) | 25.3 (19-35) | 27.7 (21.4-43) |
| Cholesterol mmol/L, mean (range) | 5.5 (3.9-8) | 5.4 (4.4-6.8) | 5.4 (3.9-8) | 5.6 (3.9-7.9) |
| Vitamin D (ng/ml), mean (range) | 28.8 (9.2-63) | 33.2 (13.3-63.9) | 30.6 (9.2-63.9) | 27.4 (8.4-78.7) |
| Smokers n | 0 | 0 | 0 | 0 |
| Previous smokers n | 4 | 2 | 6 | 2 |
| Passive smokers n | 4 | 5 | 9 | 7 |
| Fruits/veg>5: days/week, mean (range) | 5 (0-7) | 4.4 (0-7) | 4.8 (0-7) | 3.9 (0-7) |
| EDSS, median (range) | 2 (1-3.5) | 4.2 (1-8) | 2.9 (1-8) | NA |

### MRI results of pwMS and Controls

Mean lesion volumes (mm^3^) were significantly greater (p<0.001) in pwMS (10262 ± 10294) than in Controls (768 ± 1001), i.e. the mean WML volumes of pwMS were 10.2 ml vs 0.7 ml in the Controls. None of the Controls had neurological symptoms; however, SAMSEG software identified WMLs in some Control participants. Radiologist reports confirmed the presence of age-appropriate white matter hyperintensities that were identified as WMLs by the software. There was a trend for a negative association between WM hyperintensities and vitamin D concentrations in the Controls (r = −0.44; p=0.058).

In the 22 pwMS, associations were assessed between WML volumes and age, disease duration (years with MS) and disability (EDSS). Of the 22 pwMS, 10 were on disease-modifying treatment (DMT), all of whom were prescribed Interferon-β. WML volumes were associated significantly with the EDSS (p<0.02), but not with age, disease duration or DMT use (Table 3). Of the 22 pwMS, 13 had followed the PSGT Program for more than 10 years, while 9 had not followed the Program. There was a significant difference in WML volumes between the two groups (Fig 2); 4950 ± 5303 vs 17934 ± 11139 (p= 0.002).

**Table 3.** Associations of WML volumes with age, disease duration, disability and medication use in pwMS.

| Variable | Pearson correlation | P-value |
| --- | --- | --- |
| Age at MRI | 0.21 | 0.350 |
| Disease duration | 0.08 | 0.709 |
| EDSS | 0.73 | <0.02 |
| DMT use |  | 0.70* |

\* ANOVA test for equal means

**Figure 2.**
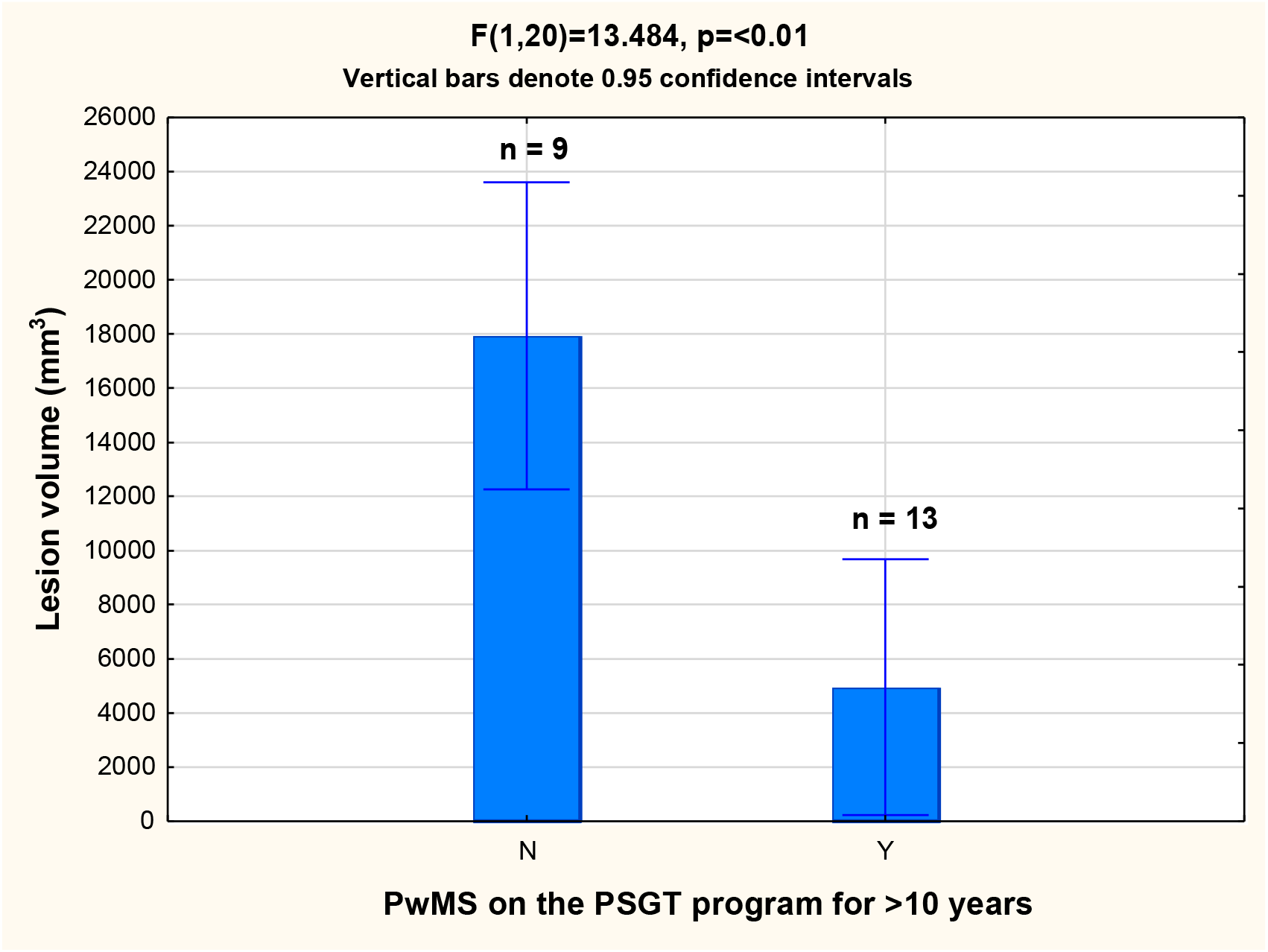
Significant difference between white matter lesion volumes of pwMS (n=13) who had followed the PSGT Program for > 10 years and pwMS (n=9) who had not, 4950 ± 5303 vs 17934 ± 11139 (p=0.002). N = no, Y = yes.

EDSS values in the pwMS who had followed the Program were significantly (p=0.02) lower (mean 2.0, range 1-3.5) than the pwMS who had not (mean 4.2, range 1-8) (Table 2); i.e. pwMS on the Program had EDSS values in the benign range after more than 10 years. They were all freely ambulatory and did not have major MS symptoms. Of the pwMS who had a disease duration of more than 10 years, and had not followed the Program (n=4), two had to use a cane for walking and 2 were wheelchair bound. The other 5 pwMS not following the Program had a disease duration of less than 10 years.

## Discussion

The present study showed that the pwMS who had followed the PSGT Program for longer than 10 years, had WML volumes that were 3 times smaller than the pwMS who had not followed the Program, a significant difference (p=0.002), despite the small study sample. Furthermore, the pwMS who had followed the Program had a favourable disability outcome after more than 10 years. If this result could be repeated in a larger sample, it would confirm our hypothesis that oligodendrocyte death and subsequent WML formation could be delayed and severe disease prevented by mitigation of risk factors, optimal nutrition and adequate blood flow to the CNS. We have previously published a case series of pwMS who have followed our Program who have experienced favourable disability outcomes after more than 10-20 years; one is an ultramarathon athlete, and one experienced a reversal of disability from EDSS 7.5 to 2.0.^5^

The rationale for including specific risk factors into the PSGT Program was gleaned from the literature and our own research experience:

1. Iron and vitamin B12-folate methylation capacity: Oligodendrocytes are extremely energy dependent, because they have to produce and maintain an enormous amount of myelin, including cholesterol that is not obtained from the circulation but has to be synthesized by the oligodendrocytes themselves.^22^ Sufficient iron and vitamin B12-folate methylation capacity is required by the mitochondria for this excessive energy generation.^6,23^ If the oligodendrocytes experience energy blackouts they perish.^6,24^
2. Vitamin D deficiency is a causal risk factor for MS^25^ since it is required to modulate the activated immune system in MS.^26^
3. Epstein-Barr virus infection has also been shown to be a causal factor for MS,^25^ and the only currently known protection available against this insult is sufficient vitamin D intake, which may need to be as high as 20,000 IU/day for short time periods.^27^ Optimal serum values for vitamin D in MS are suggested to be 30-100 nl/ml.^27^
4. Oxidation and inflammation are extremely prevalent in MS and need to be constantly counteracted by adequate intake of antioxidants; daily intake of more than 5 fruits/vegetables is inversely associated with disability.^9^ OPCs are particularly vulnerable to oxidation.^28^ If OPCs are subjected to the same risk factors as the oligodendrocytes that they are meant to replace, they may also not survive.
5. Unsaturated oils (olive oil, omega-3 and evening primrose oil) also provide protection against inflammation.^5^
6. Environmental toxins such as smoking and pesticides aggravate relapses and disability.^5,9^
7. Saturated fat intake, which contributes to obesity, was the first environmental causative risk factor for MS to be identified by Swank et al.^12^ Swank and Goodwin postulated that saturated fat impedes nourishment of tissues due to rigid chylomicra aggregates blocking the small blood vessels in the CNS, in contrast to smaller, less rigid aggregates formed from unsaturated oils in the diet.^29^
8. Avoiding food allergens reduces fatigue and improves QOL in pwMS who are susceptible to these food sensitivities.^14^
9. Stress: pwMS participating in our Program have stated that stress is an aggravating risk factor for MS and MS relapses (Gknowmix questionnaire entries, unpublished results) while exercise improves blood flow and alleviates stress and pain.^5^

In the present study, we used the freely-available software package SAMSEG ^30^ to quantify WMLs from MRI data. SAMSEG takes multi-contrast MRI data as input, regardless of scanner or pulse sequence used and segments 41 brain structures as well as WMLs using a generative model.^20^ We used both T1 and FLAIR images as input, as SAMSEG benefits from the T1 contrast for segmenting cortical gray-white matter boundaries and from the FLAIR contrast for optimum delineation of lesions. Apart from co-registering the FLAIR and T1 images and reslicing them to the same resolution, SAMSEG requires no image preprocessing. Although manual labelling has been shown to be slightly more accurate,^20^ automated methods eliminate intra- and inter-rater discrepancies. Automated tools that can quickly and reliably characterise the shape and size of WMLs are of great value for tracking disease progression and evaluating treatment efficacy from MRI data.

### Limitations

The greatest limitation of the study is the small sample size, the observational nature of the PSGT Program and the current absence of RCTs. The strength of the study is the significantly smaller WML volumes and benign disease outcomes in pwMS who had followed the PSGT Program for more than 10 years compared to those who had not (Figure 2). A limiting factor of the WML volume measurement may be the requirement in automated methods to specify a probability threshold for lesion identification. Too low a threshold may result in false positives and too high a threshold may impede sensitivity. However, since our data was acquired with a standardized protocol on the same scanner, a suboptimal threshold selection would result in a systematic over- or underestimation of lesion volume across subjects.

In conclusion, this pilot case-controlled study may provide a contribution to impact prevention and management of MS. The tentative findings suggest that WML formation could be delayed and severe disease prevented by mitigation of actionable risk factors through a diet- and lifestyle Program that protects oligodendrocytes and stimulates OPCs to remyelinate the axons during MS relapses. It is noteworthy that neuropathological studies have not identified T-cells or B-cell derived plasma cells as the main players in early lesions in MS. This suggests that oligodendrocyte death due to pathogenic environmental factors such as ischaemia, oxidative stress or viral infections, acting in concert with genetic predisposition and suboptimal dietary habits, may be the root cause of MS with immune activation as a consequence. If this were the case, different treatment strategies for MS need to be investigated. This study emphasizes the value of neuropathological- and MRI methodology to elucidate the underlying cause(s) of MS.

## Data Availability

The anonymised data set used and analyzed during this study is available from the corresponding author upon reasonable request.

## Acknowledgements

The authors thank all participants in the MS Project. We gratefully acknowledge a donation towards the MS project from a person diagnosed with MS and her family. They asked their wedding guests to donate towards the project in lieu of floral arrangements and wedding gifts; since joining the project she has been without MS symptoms for more than 15 years. We dedicate this article to the memory of her father who recently passed away and who was the Secretary of the MS Care Trust South Africa. We also acknowledge the MS Care Trust and the Cape Peninsula University of Technology, Cape Town, South Africa for their support.

## Funding

This study was funded by the National Health Laboratory Service Award Number 94139 and the South African BioDesign Initiative of the Department of Science and Innovation and the Technology Innovation Agency, South Africa (grant no. 401/01). MC Kemp was supported by Winetech, South Africa (grant no. N07/09/203). M Jaftha is funded by the Health and Welfare Sector Education and Training Authority (HW SETA) of South Africa (bursary code N716 02). The funding bodies had no role in the design of the study, collection, analysis and interpretation of data or in writing of the manuscript.

No writing assistance was utilized in the production of this manuscript.

## References

1. Barnett MH, Prineas JW. Relapsing and remitting multiple sclerosis: pathology of the newly forming lesion. Ann Neurol 2004; 55: 458–468.

2. Prineas J, Lee S. Microglia subtypes in acute, subacute, and chronic multiple sclerosis. Journal of Neuropathology and Experimental Neurology 2023; 82(8), 674–694.

3. Bergles DE, Richardson WD. Oligodendrocyte Development and Plasticity. Cold Spring Harb Perspect Biol 2015; 8(2): a020453.

4. Rooney RN, Kotze MJ, de Villiers JN, et al. Multiple sclerosis, porphyria-like symptoms, and a history of iron deficiency anemia in a family of Scottish descent. Am J Med Genet 1999; 86(2): 194–196.

5. Johannes C, Moremi KE, Kemp MC, et al. Pathology-supported genetic testing presents opportunities for improved disability outcomes in multiple sclerosis. Per Med 2023; 20(2): 107–130.

6. van Rensburg SJ, van Toorn R, Erasmus RT, et al. Pathology-supported genetic testing as a method for disability prevention in multiple sclerosis (MS). Part I. Targeting a metabolic model rather than autoimmunity. Metab Brain Dis 2021; 36(6): 1151–1167.

7. van Rensburg SJ, Hattingh C, Johannes C, et al. Pathology-supported genetic testing as a method for disability prevention in multiple sclerosis (MS). Part II. Insights from two MS cases. Metab Brain Dis 2021; 36(6): 1169–1181.

8. Kotze MJ, de Villiers JN, Warnich L, Schmidt S, et al. Lack of clinical manifestation of hereditary haemochromatosis in South African patients with multiple sclerosis. Metab Brain Dis 2006; 21(2-3): 109–120.

9. Davis W, van Rensburg SJ, Cronje FJ, et al. The fat mass and obesity-associated FTO rs9939609 polymorphism is associated with elevated homocysteine levels in patients with multiple sclerosis screened for vascular risk factors. Metab Brain Dis 2014; 29(2): 409–419.

10. Kemp MC, Johannes C, van Rensburg SJ, et al. Disability in multiple sclerosis is associated with vascular factors: An ultrasound study. J Med Imaging Radiat Sci. 2023; 54(2): 247–256.

11. Martinez Sosa S, Smith KJ. Understanding a role for hypoxia in lesion formation and location in the deep and periventricular white matter in small vessel disease and multiple sclerosis. Clin Sci (Lond) 2017; 131(20): 2503–2524.

12. Swank RL, Grimsgaard A. Multiple sclerosis: the lipid relationship. Am J Clin Nutr 1988; 48(6): 1387–1393.

13. Snetselaar LG, Cheek JJ, Fox SS, et al. Efficacy of Diet on Fatigue and Quality of Life in Multiple Sclerosis: A Systematic Review and Network Meta-analysis of Randomized Trials. Neurology 2023;100(4): e357–e366.

14. Wahls TL, Titcomb TJ, Bisht B, et al. Impact of the Swank and Wahls elimination dietary interventions on fatigue and quality of life in relapsing-remitting multiple sclerosis: The WAVES randomized parallel-arm clinical trial. Mult Scler J Exp Transl Clin 2021; 7(3): 20552173211035399. eCollection 2021 Jul-Sep.

15. Katz Sand I, Benn EKT, Fabian M, et al. Randomized-controlled trial of a modified Mediterranean dietary program for multiple sclerosis: A pilot study. Mult Scler Relat Disord 2019; 36: 101403.

16. Yadav V, Marracci G, Kim E, et al. Low-fat, plant-based diet in multiple sclerosis: A randomized controlled trial. Mult Scler Relat Disord 2016; 9:80–90.

17. Saul AM, Taylor BV, Blizzard L, Simpson-Yap S et al; Ausimmune/AusLong Investigators. A pro-inflammatory diet in people with multiple sclerosis is associated with an increased rate of relapse and increased FLAIR lesion volume on MRI in early multiple sclerosis: A prospective cohort study. Mult Scler 2023; 29(8): 1012–1023.

18. McDonald WI, Compston A, Edan G et al. Recommended diagnostic criteria for multiple sclerosis: guidelines from the International Panel on the diagnosis of multiple sclerosis. Ann Neurol 2001; 50(1): 121–127.

19. Kurtzke JF. Rating neurologic impairment in multiple sclerosis: an expanded disability status scale (EDSS). Neurology 1983; 33: 1444–1452.

20. Cerri S, Puonti O, Meier DS, et al. A Contrast-Adaptive Method for Simultaneous Whole-Brain and Lesion Segmentation in Multiple Sclerosis. NeuroImage 2021; 225: 117471.

21. World Medical Association. Declaration of Helsinki: Ethical Principles for Medical Research Involving Human Subjects. JAMA 2013; 310(20): 2191–2194.

22. Connor JR, Menzies SL. Relationship of iron to oligodendrocytes and myelination. Glia 1996; 17: 83–93

23. van Rensburg SJ, Peeters AV, van Toorn R, et al. Identification of an iron-responsive subtype in two children diagnosed with relapsing-remitting multiple sclerosis using whole exome sequencing. Mol Genet Metab Rep 2019; 19: 100465. eCollection 2019 Jun.

24. Van Rensburg SJ, Kotze MJ, Van Toorn R. 2012. The Conundrum of Iron in Multiple Sclerosis – Time for an Individualised Approach. Metab Brain Dis 2012; 27(3): 239–253.

25. Munger K, Ascherio A. Understanding the joint effects of EBV and vitamin D in MS. Mult Scler 2013; 19(12): 1554–1555.

26. Galoppin M, Kari S, Soldati S, Pal A, et al. Full spectrum of vitamin D immunomodulation in multiple sclerosis: mechanisms and therapeutic implications. Brain Commun 2022; 4(4): fcac171. eCollection 2022.

27. Disanto G, Handel AE, Damoiseaux J, et al. Vitamin D supplementation and antibodies against the Epstein-Barr virus in multiple sclerosis patients. Mult Scler 2013; 19(12):1679–1680.

28. French HM, Reid M, Mamontov P, et al. (2009) Oxidative stress disrupts oligodendrocyte maturation. J Neurosci Res 2009; 87:3076–3087.

29. Swank RL, Goodwin JW. How saturated fats may be a causative factor in multiple sclerosis and other diseases. Nutrition 2003; 19(5): 478.

30. Van Leemput K. Samseg (cross-sectional, longitudinal, MS lesions), 2023, https://surfer.nmr.mgh.harvard.edu/fswiki/Samseg

